# A protocol for a multicentre observational study across an integrated care system to understand the effect of living with frailty on access to video consultations

**DOI:** 10.1101/2022.04.14.22270318

**Authors:** Matthew Shorthose, Nick Watts, Ben Carter, Bekah Evans, Samuel Jeynes, Sue Wensley, Philip Braude

## Abstract

Risks for digital exclusion include older age, disability, and deprivation. No literature exists examining frailty as an additional possible risk factor. Many studies have shown frailty to be a more accurate predictor of adverse outcome in healthcare than age alone. Understanding risk factors for digital exclusion may help healthcare services appropriately tailor interventions. The Healthier Together integrated care system (HTICS) in Bristol is developing video consultations for clinics. The aim of this prospective observational cross-sectional study is to identify whether living with frailty is a risk factor of digital exclusion from video consultation. Data will be collected using a structured survey including patients from three sites across the Heal secondary care services at North Bristol Trust, and then two subsequent sites will be included across HTICS. Data will be collected on known risk factors for digital exclusion, frailty using the Clinical Frailty Scale, and will cover four domains for digital exclusion as defined by NHS Digital: access, skill, motivation, and confidence. Data will be presented with descriptive statistics. The association between frailty and outcomes will be analysed with a multivariable logistic regression.

**Key Study Contacts:** 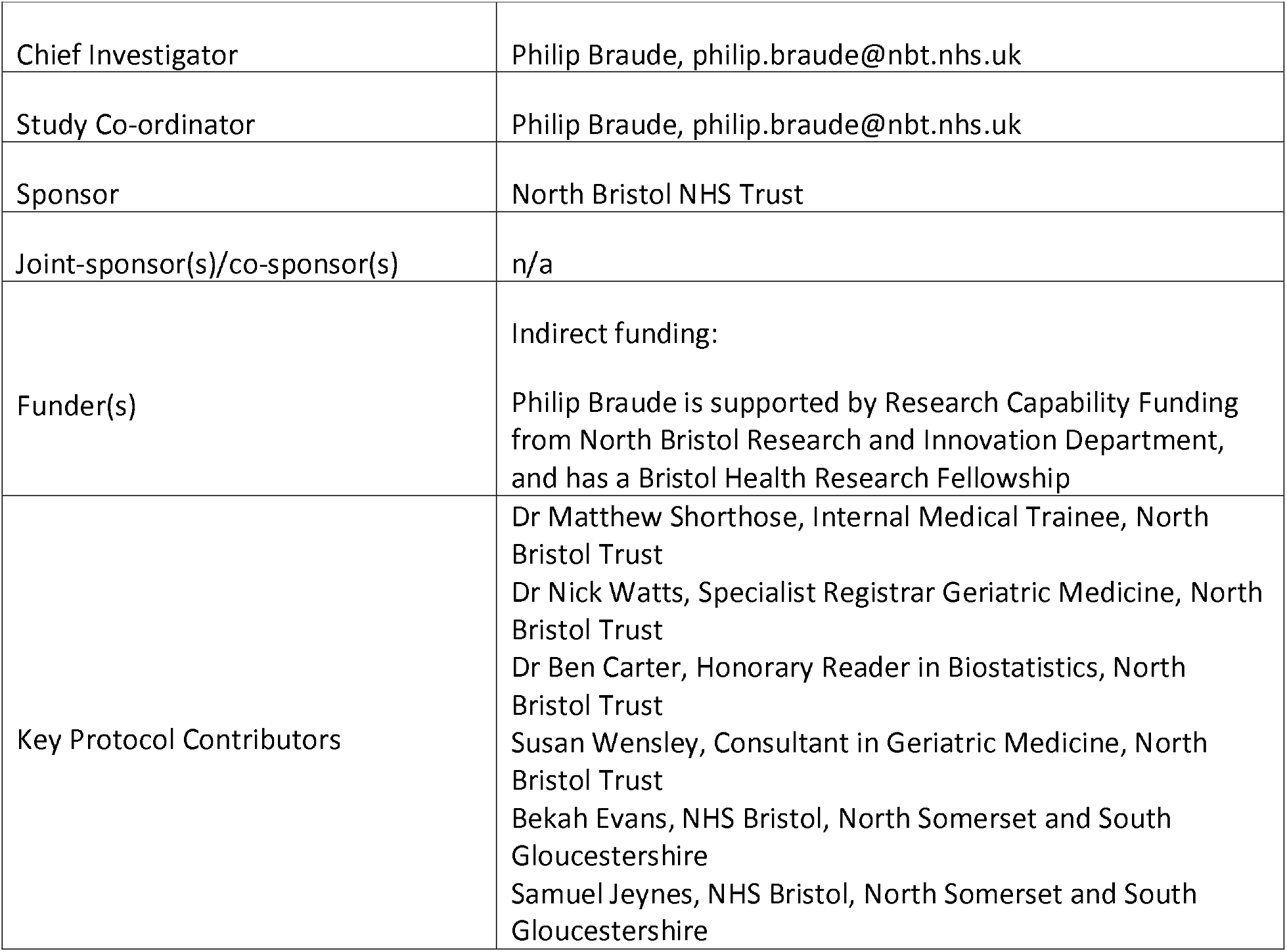

**Study Summary:** 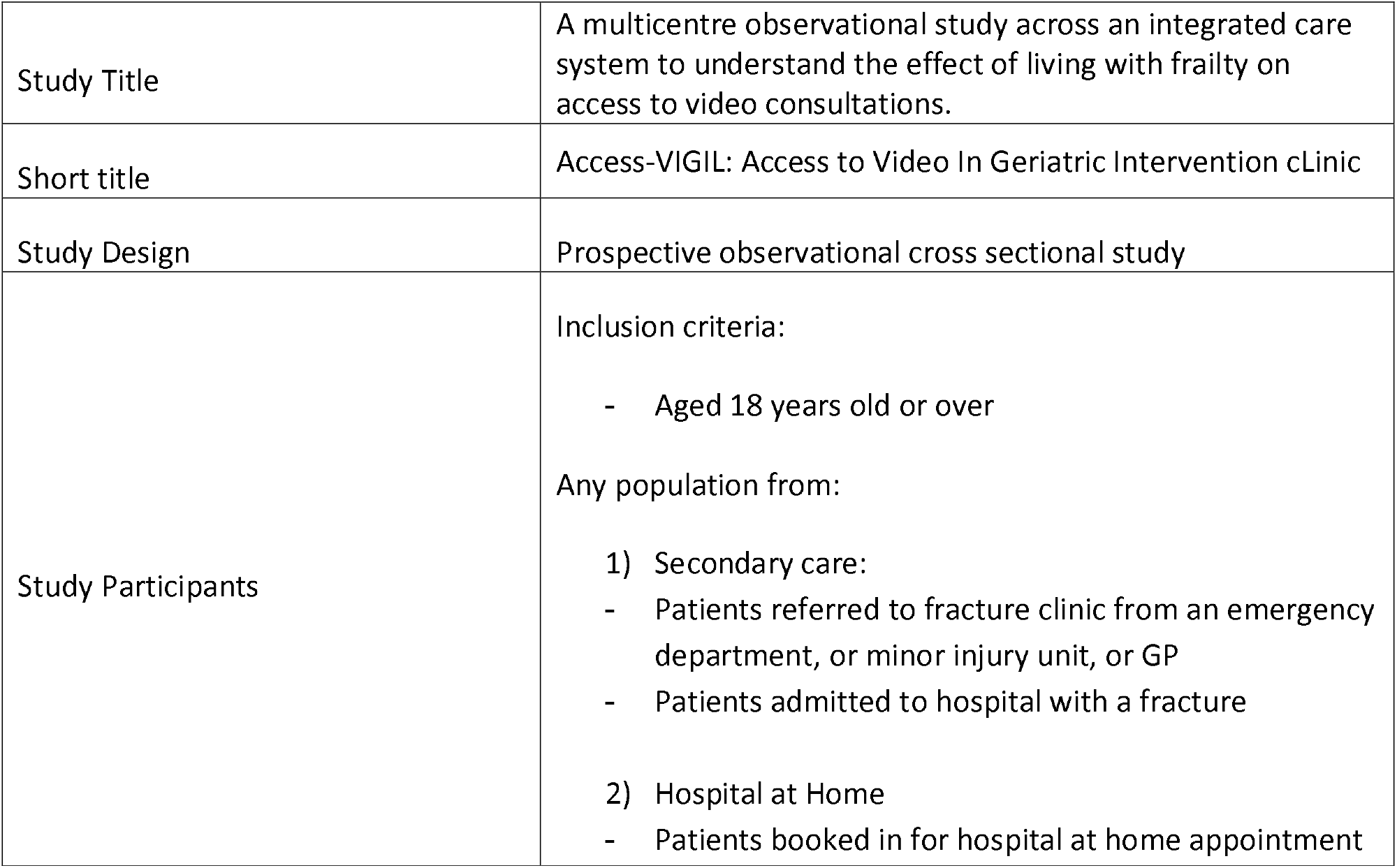

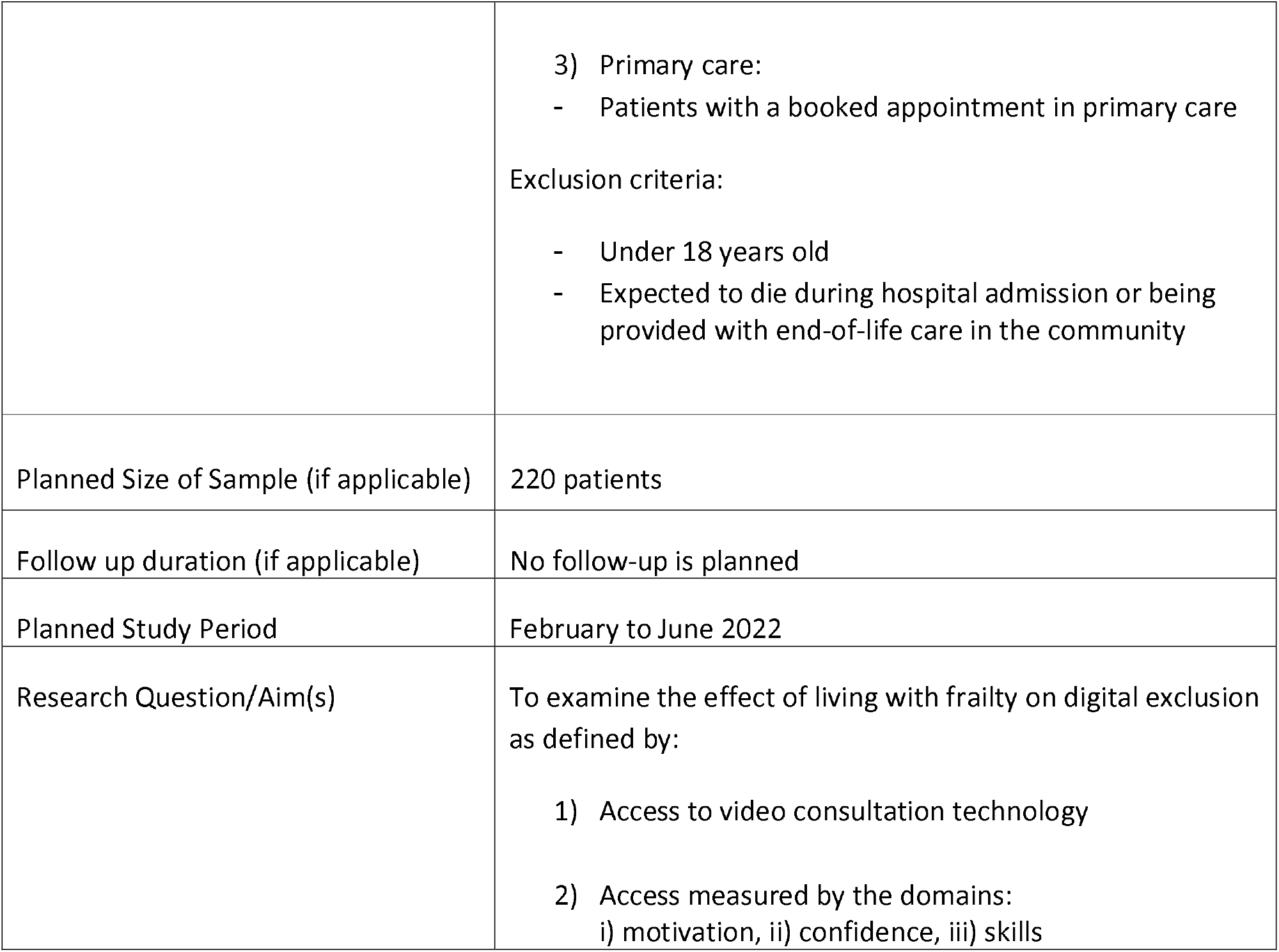

**Funding and Support in Kind:** 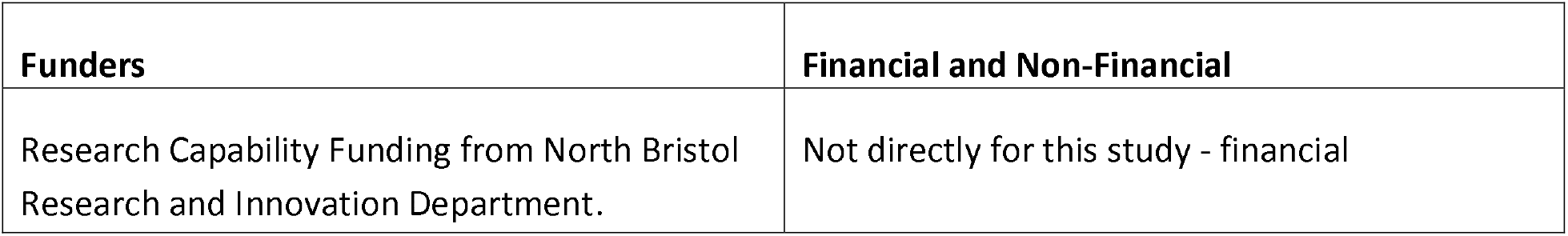

**Role of Study Sponsor and Funder:** The pilot at North Bristol Trust (NBT) is overseen by Neuromusculoskeletal (NMSK) Division governance. NBT NMSK Division takes responsibility for ensuring that the design of the study meets appropriate standards and that arrangements are in place to ensure appropriate conduct and reporting. Governance oversight for Hospital at Home was registered with NBT NMSK, and for primary care with Horfield Health Centre.

**Roles and Responsibilities:** 

**Study Flow Chart:** 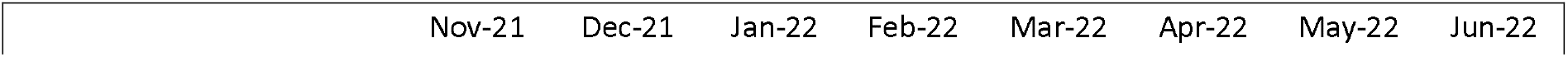

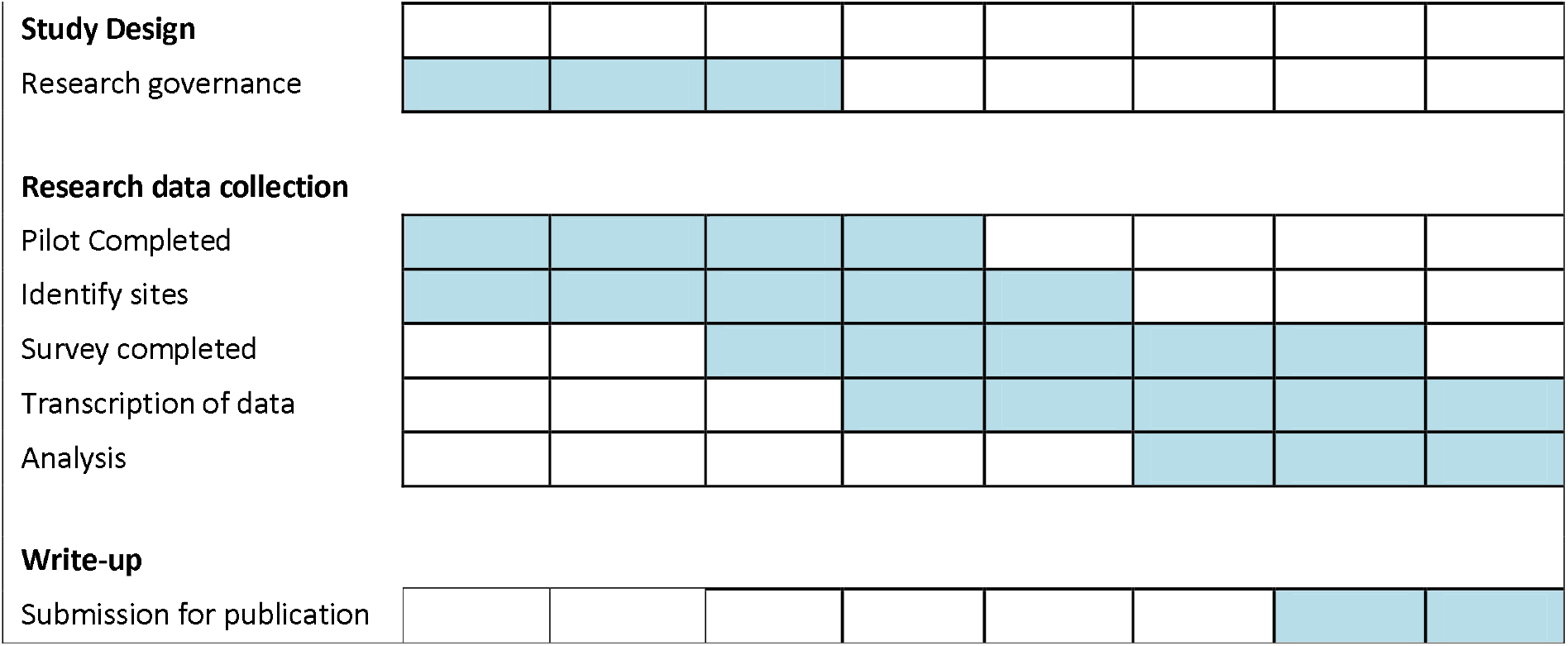

## 1 Background

Since the beginning of the COVID-19 pandemic, services have increasingly been held via video or telephone (1). Even prior to the pandemic the NHS Long Term Plan and the Royal College of Physicians were calling for alternatives to face-to-face consultations (2). For this to be a long term successful strategy patients should not be excluded from accessing these resources(3,4). As part of this move towards increased used of digital resources, we should aim to offer the right type of clinic to the right people and to identify the kind of help needed to access video clinics.

Risk factors for digital exclusion include certain ethnic groups, deprivation, and older age. However, many older people have excellent access to digital technology (5). One risk factor that has not been explored is living with frailty. Frailty is a state of reduced physiological and cognitive reserve to cope with a stressor. It is often associated with other geriatric syndromes such as cognitive and physical deterioration. The Clinical Frailty Scale (CFS) is a tool which was originally designed as a way of summarizing the overall fitness or frailty of an older adult (6). It gives a score of 1-9 after evaluating specific domains including comorbidity, function, and cognition (7). It can be used at the start of a patient pathway to appropriately stream patients into tailored pathways of care.

Keränen et al identified an association between the 3-point study of osteoporotic fracture index and reduced computer usage in Finland, independent of age (8). Liu et al examined the effect of frailty on people that had accessed a single secondary care geriatric medicine clinic by video or telephone and found frailty was a significant risk factor. However the analysis had not adjusted for other risk factors for digital exclusion such as deprivation(9). The UK government defined exclusion from digital technology using four key domains: 1) access 2) skills 3) motivation, and 4) confidence (3). No study we are aware of has examined how frailty may be associated with factors contributing to digital exclusion using these domains.

This cross-sectional observational study will aim to identify whether living with frailty is a risk factor of digital exclusion from video consultation. It will look to adjust for known risk factors for digital exclusion across an integrated care system.

## 2 Research Question

### 2.1 Objectives

The aim of this study is to examine the effect of living with frailty on digital exclusion. The study aims were defined by access to video consultation as measured by objectives:

a. Technology access
b. Skills in video consultation
c. Confidence in video consultation
d. Motivation and experience of internet usage

### 2.2 Primary outcome

The primary objective of being digital excluded from video consultation will be defined as no access to a computer with webcam, smartphone with camera, or tablet with camera.

### 2.3 Secondary objectives

Secondary objectives are to characterize frailty amongst the sample and assess the association between frailty and:

a. Skills in video consultation
b. Confidence in video consultation
c. Motivation and experience of internet usage

## 3 Study Design and Methods of Data Collection

### 3.1 Study design

A cross-sectional observational study will be undertaken. Sites at North Bristol Trust with additional sites identified subsequently across the integrated care system. A structured survey will be used designed by key stakeholders using questions from a question bank on digital technology (10).

### 3.2 Data Collection

Data will be collected using a semi-structured survey either in person or by telephone.

Basic demographic variables will be taken from patient’s notes, if possible, except for Clinical Frailty Score which will be determined by the interviewer at the beginning of the survey. If a variable is not available from a patient’s notes, it will be asked at the beginning of the survey. Each interviewer will have completed online training regarding the Clinical Frailty Score prior to conducting interviews.

In testing, survey completion takes four minutes. The survey will be conducted by four methods depending on the population, each using a standardized case reporting form:

1. for patients admitted – data collection in person
2. for patients discharged – in person at clinic or by telephone / video
3. for patients in interface care – in person at clinic or by telephone / video
4. for patients in primary care – in person while at the surgery or by telephone / video

### 3.3 Variables

#### 3.3.1 Clinical contextual variables collected to describe the sample

Basic demographics to be included: age, sex (male, female), frailty (Clinical Frailty Scale 1-9), ethnicity (using Gov.uk list(11)), index of multiple deprivation, co-morbidity (Charlson comorbidity index (CCI)), living situation (house, flat, assisted living, care home), memory problems Sensory impairment – reading difficulty (considerable, moderate, no); hearing difficulty (considerable, moderate, no); memory impairment (yes, no); cognitive diagnosis.

#### 3.3.2 Primary Outcome

The primary outcome of digital exclusion which is no access to video technology. No access to a device with enabled technology to conduct a videocall is defined as access to a computer, smartphone, or tablet. This could be their own, or someone else’s that they may use for the videocall.

#### 3.3.2 Secondary analysis of the Primary Outcome

The primary outcome will be re-analysed, but after defining access to a device which is owned by the patient and excludes any technology shared to the patient.

#### 3.3.3 Secondary Outcomes

##### Technology Skill

Patients were defined having the necessary technology skill if they had taken part in video call in the last year which they had set up, or rated their ability as fair or better.

##### Confidence Using Technology

Patient confidence was self-reported as a patient being quite or very confident in participating in a video call.

##### Motivation to use technology

Motivation to use online technology was defined as using any online tool including (social communication, social media, entertainment, personal finance, online shopping, apps for health, etc)

##### Technology usage

Motivation to undertake a videocall is defined using typical internet usage. If a patient is self-reported as using the internet weekly (or more regularly) they are defined as frequent users. Patients with usage of within 3 months, or less frequent are described as non-frequent users and lacked motivation to use technology.

#### 3.3.5 Primary Exposure

Frailty is the main exposure of this investigation. It will be measured using the Clinical Frailty Scale (6). This is a 9 point scale from CFS=1 a very fit patient, to CFS=8 very severely frail, CFS=9 is a patient that is terminally unwell with an estimated survival of less than 6 months. CFS 1 to 3 describe patients that are not frail. CFS=4 describes a patient that is very mildly frail. CFS=5 describes a mildly frail patient, and CFS=6 a moderately frail patient, and CFS=7-8 were severely frail patients.

The CFS has been validated to be recorded in person, as well as over the telephone following a structured set of questions or using a smartphone application (12).

#### 3.3 Data Storage

Data will be recorded either directly into a bespoke spreadsheet, or on a standardized case reporting paper form and transcribed later.

Electronic data will be stored securely at the local site. Access to the data will be by the study team only through a password protected file. Paper case reporting forms prior to transcription to the electronic database will be stored in a locked area at each site.

Data will be kept for 3 years after the study closes in of case external requests to verify published material. Participants’ contact details will be stored separately from the data entry spreadsheet. A unique key will be used to identify patients via a study ID number only.

## 4 Study Setting

Three sites across the Healthier Together integrated care system will be included. Healthier Together is a partnership across Bristol, North Somerset and South Gloucestershire with the aim of providing joined up services across the population.

The pilot site for the study will be the secondary care services delivered for acute fractures in North Bristol Trust, which is the largest hospital in South West England. The two subsequent sites will be an interface care setting and a primary care setting. The interface care will be North Bristol Trust Hospital at Home service which delivers acute healthcare to patients in the comfort of their own home as an alternative to hospital admission. Treatments include administration of intravenous antibiotics, blood monitoring, wound care, and welfare calls. The primary care setting will be Horfield Health Centre providing services to 16,500 patients from North Bristol including areas Lockleaze, Horfield and parts of Ashley Down, Henleaze, Filton and Northville.

## 5 Sample and Recruitment

### 5.1 Inclusion and exclusion

#### 5.1.1 Inclusion criteria

- Aged 18 years old or over

And any population from:

1. Secondary care:
  - Patients referred to fracture clinic from an emergency department, or minor injury unit, or GP
  - Patients admitted to hospital with a fracture
2. Hospital at Home
  - Patients booked in for Hospital at Home appointment
3. Primary care:
  Patients with a booked appointment in general practice

#### 5.1.2 Exclusion criteria

- Under 18 years old
- Expected to die during hospital admission or being provided with end-of-life care in the community

### 5.2 Sampling

#### 5.2.1 Size of sample

Liu et al found an ∼40% reduction in odds in those living with frailty having digital access. Using Liu we have assumed that 80% of those not living with frailty and 60% of those that are living with frailty will have digital access(9). Using a Chi-squared test with 90% power and with a 5% significance level to detect this difference 220 patients would need to be enrolled into the study.

#### 5.2.2 Sampling technique

Data will be collected prospectively. Convenience sampling will be used. Patients will be contacted during the study period until the sample size has been reached. A target sample population of 100 patients in each setting will be set.

### 6.3 Recruitment

#### 6.3.1 Sample identification

##### Healthier Together ICS

- North Bristol Trust: Patients will be identified prospectively over one week through referral to fracture clinic from Southmead Emergency Department, Yate Minor Injuries Unit, or their GP. Patients admitted to Southmead Hospital will be identified by the inpatient major trauma team and by the orthogeriatric team as part of their normal referral system and inpatient management. Preliminary data showed around 30 patients would be seen per a day.
- North Bristol Hospital at Home: Patients will be identified retrospectively over 3 months, with data collected prospectively, from those booked into the hospital at home team at North Bristol Trust. Preliminary data showed 30 patients would be seen per month.
- Horfield Health Centre: Patients will be identified prospectively over one week by those booked into an appointment at their local clinic. Preliminary data showed 150 patients would be seen per day.

#### 6.3.2 Consent

Consent will not be required given this study is being conducted as part of a service evaluation.

## 7 Statistical Considerations

This protocol and statistical analysis plan was pre-specified using a pre-defined hypothesis before the study data was accessed. All included covariates were defined based on clinical importance and not using any machine learning methodologies or model building techniques.

### 7.1 Descriptive data

The sample demographic and clinical characteristics will be described by: digital exclusion; and frailty.

### 7.2 Data Analysis

We will use a multivariable logistic regression to fit the association between digital inclusion and CFS (1-3, 4-5, 6-8). Our primary analysis will adjust for: enrolment site (Healthier Together ICS, North Bristol Hospital, and Horfield Health Centre), age group (under 65 years old, 65-79, 80 or over), Charlson comorbidity index (1-2, 3-4, 5+), index of multiple deprivation (1-3, 4-7, 8-10)(9). We will present the adjusted Odds ratio (aOR) with 95% confidence intervals (95%CI) alongside the crude OR, and p-values. All analyses will be carried out within Stata v17 (or later).

### 7.3 Data completeness

Missing data will be explored for pattern missingness and missing item data will be presented for each covariate to describe the sample. If missing data is suspected to be associated with any clinical characteristics the data analysis may be revised to account for the missingness or additional sensitivity analyses carried out to assess the primary analysis under a range of potential scenarios.

### 7.4 Subgroup Analyses

The primary outcome comparing the aOR for those frails (CFS 1-3) versus not frail (CFS 4-8) will be analysed in the following subgroups: age, Charlson comorbidity Index, Index of Multiple deprivation, living situation, and memory problems

## 8. Ethical and Regulatory Considerations

### 8.1 Assessment and management of risk

It is not anticipated that the survey will cause any distress or problems to patients. However, it can be anticipated that certain risks in contacting patients can be mitigated:

1. If patients have questions as to their clinical follow-up they will be advised they will hear from the clinic with a time, or we will be able to give them the time and date if it has already been arranged.
2. If patients have medical questions as to their current management, they will be directed to either contact their GP, return to the emergency department, or to call the number on their appointment letter.

### 8.2 Quality improvement and ethical approvals

Three service evaluations have been brought together under the banner of one cross-sectional study. All three services are using or have aspirations to develop video consultation services. Given the service evaluation nature of the study it has been registered with the Quality and Safety Improvement Team of North Bristol NHS Trust. It was deemed in discussion with the lead for the Quality and Safety Improvement Team that ethical approval was not required given the data collected were to be directly used to evaluate current service delivery and allow further service development. The study has been reviewed by the quality improvement leads for each site and registered locally. Approval registration IDs can be found on the front page of this protocol.

### 8.3 Amendments

Any study amendments will be submitted for review by the QI governance lead at each of the sites.

### 8.4 Peer review

The protocol has been reviewed by the Chief Investigator and study management group prior to being sent for assesssment. The QI governance lead for NMSK at North Bristol Trust has reviewed the pilot protocol. Each QI lead will review the protocl at local sites.

### 8.5 Patient Involvement

#### Patient involvement

- In development of the pilot work 67 service user’s feedback from a telephone survey has been published (13).
- The department of medicine for older people service user group was consulted
- Conversations occurred with ten patients from the user group having used the geriatric medicine clinic

### 8.6 Access to the Final Study Dataset

The anonymized data will only be available to the Chief Investigator (Philip Braude) and the study management group named in the protocol. Responses will be anonymised. Files will be password protected and stored on the hospital’s secure server, only accessible from a local service-maintained computer with employee login. Any transfer of data required will occur only using secure servers and encrypted files.

In line with many peer reviewed journal’s policy for data sharing, data sharing may be offered to third parties only on request to the study CI with review by the study sponsor to ensure legitimate academic interest. Data will be shared with anonymous records if deemed appropriate, arranged via data sharing agreement, and transferred using secure systems. If requests originate from outside the EU this will be discussed with the study sponsor.

## 9 Dissemination Policy

### 9.1 Dissemination policy

The data will be owned by North Bristol Trust as the sponsor. On completion of the study the data will be analysed and a study report completed.

It will be disseminated locally via the trust operational update and events, as well as more widely through national and international conference presentations. A summary of the work will be submitted for peer-reviewed publications.

### 9.2 Authorship Eligibility Guidelines

Authorship will take into account all persons involved in the study design, analysis and write up. This will be accurately reflected when any papers are submitted for peer-reviewed publication.

## 10 Changes to the Protocol

Any changes to the protocol prior to data collection will following a study team review. No changes to the protocol will be made following the completion of data collection. Changes at this stage will be reported in the primary analysis paper under the section “Changes from the published protocol”.

## Supporting information

HRA decision tool

Questionnaire for survey

## Data Availability

All data produced in the present study are available upon reasonable request to the authors

