## Supplementary material for "A protocol for a multicentre observational study across an integrated care system to understand the effect of living with frailty on access to video consultations": HRA decision tool

Go straight to content.

Medical Research Council

NHS Health Research Authority

### Is my study research?

**To print your result with title and IRAS Project ID please enter your details below:**

Title of your research:

A multicentre observational study across an integrated care system to understand the effect of living with frailty on access to video consultations

IRAS Project ID (if available):

You selected:

- **'No'** - Are the participants in your study randomised to different groups?
- **'No'** - Does your study protocol demand changing treatment/ patient care from accepted standards for any of the patients involved?
- **'No'** - Are your findings going to be generalisable?

**Your study would NOT be considered Research by the NHS.**

You may still need other approvals.

Researchers requiring further advice (e.g. those not confident with the outcome of this tool) should contact their R&D office or sponsor in the first instance, or the **HRA** to discuss your study. If contacting the HRA for advice, do this by sending an outline of the project (maximum one page), summarising its purpose, methodology, type of participant and planned location as well as a copy of this results page and a summary of the aspects of the decision(s) that you need further advice on to the HRA Queries Line at ****.

For more information please visit the **Defining Research** table.

**Follow this link to start again.**

Print This Page

NOTE: If using Internet Explorer please use browser print function.

**About this tool** **Feedback** **Contact** **Glossary** **Accessibility**
