## Supplementary material for "A protocol for a multicentre observational study across an integrated care system to understand the effect of living with frailty on access to video consultations": Questionnaire for survey

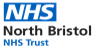

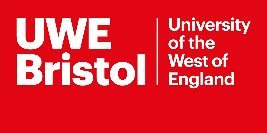

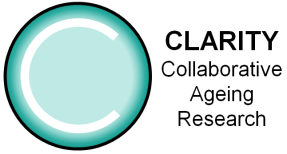

Survey to evaluate access to video consultations

We are developing video clinics for health service. We want to collect information about who can access video clinics. This information will help us offer the right type of clinic to the right people and to identify the kind of help needed to access video clinics. We are also investigating what areas of technology people already use so that we can develop additional educational resources and help.

We are surveying people who are looked after in this service. The information here will be kept confidential and only used for this project. All information will be destroyed at the end of the study. Any information that will be used in publications will be grouped and anonymized so no one can work out who answered the questions. The data will be shared anonymously with North Bristol Trust to be analysed.

If you would like to know more about this study, please contact the Chief Investigator Philip Braude via or telephone: 0117 414 6442.

| Study ID* |  |  | DOB* |  |
| --- | --- | --- | --- | --- |
|  |  |  | MRN* |  |
| **Ethnicity** | British; Irish; Any other White background; White and Black Caribbean; White and Black African; White and Asian; Any other mixed background; Indian; Pakistani; Bangladeshi; Any other Asian background; Caribbean; African; Any other Black background; Chinese; Any other ethnic group; Traveller; Arab; Not stated |  | Contact point* | In person  Phone |
|  |  |  | Sex* | Male  Female |
|  |  |  | Postcode* |  |
|  |  |  | Source* | Inpatient  Outpatient |
|  |  |  | **Clinical frailty score** |  |
|  |  |  | **Living situation** | House  Flat  Assisted living  Care home |
| **Past Medical History** | | | | |

* = can be completed before patient contact. **BOLD** = patient contact required

**Questions**

| Patient able to partake in survey | Yes | No |
| --- | --- | --- |

| Person answering questions | Patient | Relative | Carer | Other |
| --- | --- | --- | --- | --- |

1) Do you have difficulties in close sight activities such as reading?

| Yes, considerable | Yes, moderate | No |
| --- | --- | --- |

2) Do you have difficulties hearing when using the telephone or other communication devices?

| Yes, considerable | Yes, moderate | No |
| --- | --- | --- |

3) Do you have a diagnosis of any memory problems that affect your day-to-day activities?

| Yes | No |
| --- | --- |
| Diagnosis: | |

4) Devices

| **Do you have your own?** | | **If no, can you access someone else’s?** | |
| --- | --- | --- | --- |
| *Computer with webcam* | | *Computer with webcam* | |
| Yes | No | Yes | No |
| *Smartphone (phone that can show videos and has a camera)* | | *Smartphone (phone that can show videos)* | |
| Yes | No | Yes | No |
| *Tablet (with camera)* | | *Tablet* | |
| Yes | No | Yes | No |

5) In the last 12 months have you used a:

| **Computer** | | | |
| --- | --- | --- | --- |
| Yes without difficulty | Yes with difficulty | | No |
| **Smartphone** | | | |
| Yes without difficulty | Yes with difficulty | | No |
| **Tablet** | | | |
| Yes without difficulty | Yes with difficulty | No | |

| Yes – have access and use at home | No do not have access at home | Yes – have access but don’t use at home | Don’t know |
| --- | --- | --- | --- |

6) Does your home have access to the internet?
7) How often do you use the internet?

| More than once a day | Once a day | Almost every day | At least weekly | Used within the last 3 months | Between 3 months and a year ago | More than a year ago | Never used the internet |
| --- | --- | --- | --- | --- | --- | --- | --- |

8) Have you ever participated or been involved in a video call on any device?

| Yes | No – go to 12 |
| --- | --- |

9) Who set up or joined the videocall?

| You |  |  | Friend |  |  | Relative |  |  | Carer |  |  | Someone else |
| --- | --- | --- | --- | --- | --- | --- | --- | --- | --- | --- | --- | --- |

| Could you/they set up or join the videocall again? | | Yes | |  | No |
| --- | --- | --- | --- | --- | --- |

10) How would you rate your ability to participate in a videocall?

| Excellent | Good | Fair | Poor | Bad | Don’t know |
| --- | --- | --- | --- | --- | --- |

11) How would you rate your confidence participating in videocalls?

| Very confident | Quite confident | Neither confident or unconfident | Not very confident | Not at all confident | Don’t know |
| --- | --- | --- | --- | --- | --- |

Why did you rate your confidence this way?

12) Do you use any of these other online tools?

| Social communication *(such as Email, WhatsApp, Messenger)* | Yes / No |
| --- | --- |
| Social media *(Facebook, Instagram, Twitter etc)* | Yes / No |
| Entertainment (*streaming film or TV, gaming etc*) | Yes / No |
| Personal finance and banking | Yes / No |
| Online household tasks (*food shopping, organising bills*) | Yes / No |
| Apps for health (*NHS app, NHS COVID app*) | Yes / No |
| Online health related tasks or management (*e.g. fitness related, medication reminders, apps for specific conditions such a COPD or diabetes, NHS apps*) | Yes / No |
| Other (please specify) | Yes / No |
